# Predicting the Growth and Trend of COVID-19 Pandemic using Machine Learning and Cloud Computing

**DOI:** 10.1101/2020.05.06.20091900

**Authors:** Shreshth Tuli, Shikhar Tuli, Rakesh Tuli, Sukhpal Singh Gill

## Abstract

The outbreak of COVID-19 Coronavirus, namely SARS-CoV-2, has created a calamitous situation throughout the world. The cumulative incidence of COVID-19 is rapidly increasing day by day. Machine Learning (ML) and Cloud Computing can be deployed very effectively to track the disease, predict growth of the epidemic and design strategies and policy to manage its spread. This study applies an improved mathematical model to analyse and predict the growth of the epidemic. An ML-based improved model has been applied to predict the potential threat of COVID-19 in countries worldwide. We show that using iterative weighting for fitting Generalized Inverse Weibull distribution, a better fit can be obtained to develop a prediction framework. This can be deployed on a cloud computing platform for more accurate and real-time prediction of the growth behavior of the epidemic. A data driven approach with higher accuracy as here can be very useful for a proactive response from the government and citizens. Finally, we propose a set of research opportunities and setup grounds for further practical applications. Predicted curves for some of the most affected countries can be seen at https://collaboration.coraltele.com/covid/.

## 1 Introduction

The novel Coronavirus disease (COVID-19) was first reported on 31 December 2019 in the Wuhan, Hubei Province, China. It started spreading rapidly across the world [1]. The cumulative incidence of the causitive virus (SARS-CoV-2) is rapidly increasing and has affected 196 countries and territories with USA, Spain, Italy, U.K. and France being the most affected [2]. World Health Organization (WHO) has declared the coronavirus outbreak a pandemic, while the virus continues to spread [3]. As on 4 May 2020, a total of 3,581,884 confirmed positive cases have been reported leading to 248,558 deaths [2]. The major difference between the pandemic caused by CoV-2 and related viruses, like SARS and MERS is the ability of CoV-2 to spread rapidly through human contact and leave nearly 20% infected subjects as symptom-less carriers [4]. Moreover, various studies reported that the disease caused by CoV-2 is more dangerous for people with weak immune system. The elderly people and patients with life threatening diseases like cancer, diabetes, neurological conditions, coronary heart disease and HIV/AIDS are more vulnerable to severe effects of COVID-19 [5]. In the absence of any curative drug, the only solution is to slow down the spread by exercising “social distancing” to block the chain of spread of the virus. This behavior of CoV-2 requires developing robust mathematical basis for tracking its spread and automation of the tracking tools for on line dynamic decision making.

There is a need for innovative solutions to develop, manage and analyse big data on the growing network of infected subjects, patient details, their community movements, and integrate with clinical trials and, pharmaceutical, genomic and public health data [6]. Multiple sources of data including, text messages, online communications, social media and web articles can be very helpful in analyzing the growth of infection with community behaviour. Wrapping this data with ML and Artificial Intelligence (AI), researchers can forecast where and when, the disease is likely to spread, and notify those regions to match the required arrangements. Travel history of infected subjects can be tracked automatically, to study epidemiological correlations with the spread of the disease. Some community transmission based effects have been studied in other works (https://www.cdc.gov/mmwr/volumes/69/wr/mm6915e1.htm). Infrastructure for the storage and analytics of such huge data for further processing needs to be developed in an efficient and cost-effective manner. This needs to be organized through utilization of cloud computing and AI solutions [7]. Alibaba developed cloud and AI solutions to help China, fight against coronavirus, predict the peak, size and duration of the outbreak, which is claimed to have been implemented with 98% accuracy in real world tests in various regions of China [8]. FDifferent types of pneumonia can be resolved using ML-based CT Image Analytics Solution, which can be helpful to monitor the patients with COVID-19 [9]. Details can be seen at https://spectrum.ieee.org/the-human-os/biomedical/imaging/hospitals-deploy-ai-tools-detect-covid19-chest-scans. The development of vaccine for COVID-19 can also be accelerated by analysing the genome sequences and molecular docking, deploying various ML and AI techniques [10].

### Motivation and Our Contributions

ML [11] can be utilized to handle large data and intelligently predict the spread of the disease. Cloud computing [12] can be used to rapidly enhance the prediction process using high-speed computations [7]. Novel energy-efficient edge systems can be used to procure data, in order to bring down power consumption. In this paper, we present a prediction model deployed using FogBus framework [13] for accurate prediction of the number of COVID-19 cases, the rise and the fall of the number of cases in near future and the date when various countries may expect the pandemic to end. We also provide a detailed comparison with a baseline model and show how catastrophic the effects can be if poorly fitting models are used. We present a prediction scheme based on the ML model, which can be used in remote cloud nodes for real-time prediction allowing governments and citizens to respond proactively. Finally, we summarize this work and present various research directions.

### Article structure

The rest of the paper is organized as follows: Section 2 presents the prediction model and performance comparison. Section 3 concludes the work and describes the future research opportunities. Section 4 provides details of open repositories for the dataset, code and results.

## 2 Prediction Model and Performance Comparison

Machine Learning (ML) and Data Science community are striving hard to improve the forecasts of epidemiological models and analyze the information flowing over Twitter for the development of management strategies, and the assessment of impact of policies to curb its spread. Various datasets in this regard have been openly released to the public. Yet, there is a need to capture, develop and analyse more data as the COVID-19 grows worldwide [14, 15].

The novel coronavirus is leaving a deep socio-economic impact globally. Due to the ease of virus transmission, primarily through droplets of saliva or discharge from the nose when an infected person coughs or sneezes, countries which are densely populated need to be on a higher alert [16]. To gain more insight on how COVID-19 is impacting the world population and to predict the number of COVID-19 cases and dates when the pandemic may be expected to end in various countries, we propose a Machine Learning model that can be run continuously on Cloud Data Centers (CDCs) for accurate spread prediction and proactive development of strategic response by the government and citizens.

### Dataset

The dataset used in this case study is the Our World in Data by Hannah Ritchie.^1^ The dataset is updated daily from the World Health Organization (WHO) situation reports.^2^ More details about the dataset are available at: https://ourworldindata.org/coronavirus-source-data.

### Cloud framework

The ML models are built to make a good advanced prediction of the number of new cases and the dates when the pandemic might end. To provide fail-safe computation and quick data analysis, we propose a framework to deploy these models on cloud datacenters, as shown in Figure 1.In a cloud based environment, the government hospitals and private health-centers continuously send their positive patient count. Population density, average and median age, weather conditions, health facilities etc. are also to be integrated for enhancing the accuracy of the predictions. For this case study, we used three instances of single core *Azure B1s* virtual machines with 1-GiB RAM, SSD Storage and 64-bit Microsoft Windows Server 2016^1^. We used the HealthFog [11] framework leveraging the FogBus [13] for deploying multiple analysis tasks in an ensemble learning fashion to predict various metrics, like the number of anticipated facilities to manage patients and the hospitals. We analyzed that the cost of tracking patients on a daily basis, amortized CPU consumption and Cloud execution is 37% and only 1.2 USD per day. As the dataset size increases, computationally more powerful resources would be needed.

**Figure 1:**
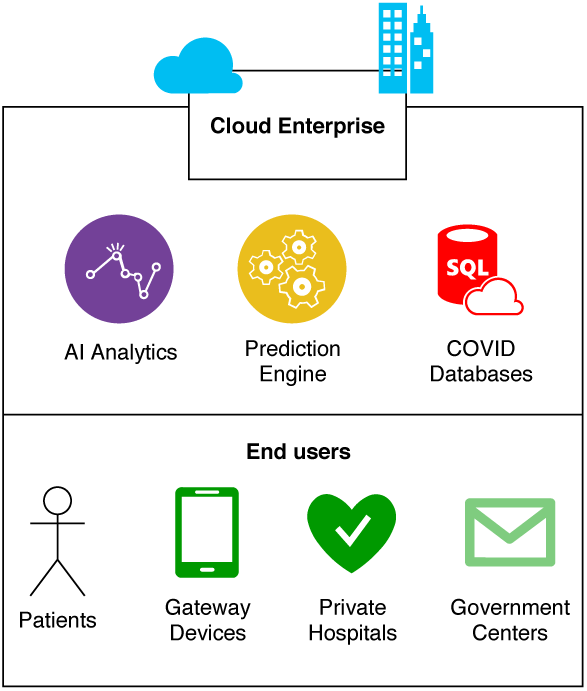
Proposed Cloud based AI framework for COVID-19 related analytics.

### ML model

Many recent works have suggested that the COVID-19 spread follows exponential distribution [17, 18, 19]. As per empirical evaluations and previous datasets on SARS-CoV-1 virus pandemic, many sources have shown that data corresponding to new cases with time has large number of outliers and may or may not follow a standard distribution like Gaussian or Exponential [20, 21, 22, 23]. In recent study by Data-Driven Innovation Laboratory, Singapore University of Technology and Design (SUTD), the regression curves were drawn using the Susceptible-Infected-Recovered model [24] and Gaussian distribution was deployed to estimate the number of cases with time. However, in the previously reported studies on the earlier version of the virus, namely SARA-CoV-1, the data was reported to follow Generalized Inverse Weibull (GIW) Distribution [25] better than Gaussian as shown in Figure 2 (details of Robust Weibull fitting follow in the next section). Detailed comparison for SARS-CoV-2 has been described in the next section. This fits the following function to the data:

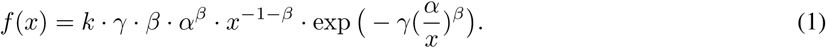

**Figure 2:**
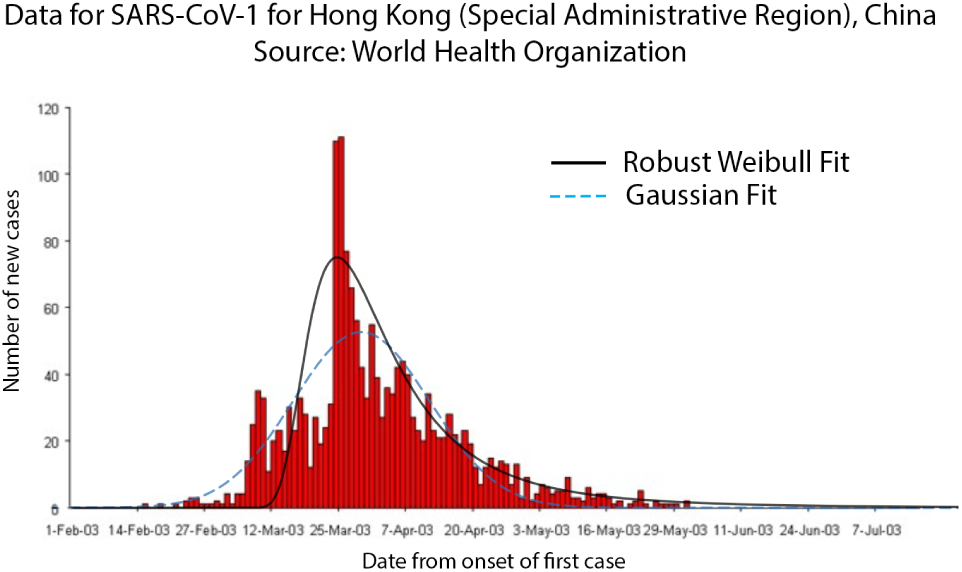
Fit curves for SARS-CoV-1 pandemic for Hong Kong (SAR), China. Data source: WHO epidemic curves (https://www.who.int/csr/sars/epicurve/epiindex/en/index4.html)

Here, *f*(*x*) denotes the number of cases with *x*, where *x* > 0 is the time in number of days from the first case, and *α, β, γ >* 0, 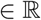 are parameters of the model. Now, we can find the appropriate values of the parameters *α, β* and *γ* to minimize the error between the predicted cases (*y = f*(*x*)) and the actual cases (*ŷ*). This can be done using the popular Machine Learning technique of Levenberg-Marquardt (LM) for curve fitting [26]. However, as various sources have suggested, in initial stages of COVID-19 the data has many outliers and noise. This makes it hard to accurately predict the number of cases. Thus, we propose an iterative weighting strategy and call our fitting technique “Robust Fitting”. A diagrammatic representation of the iterative weighting process is shown in Figure 3.

**Figure 3:**
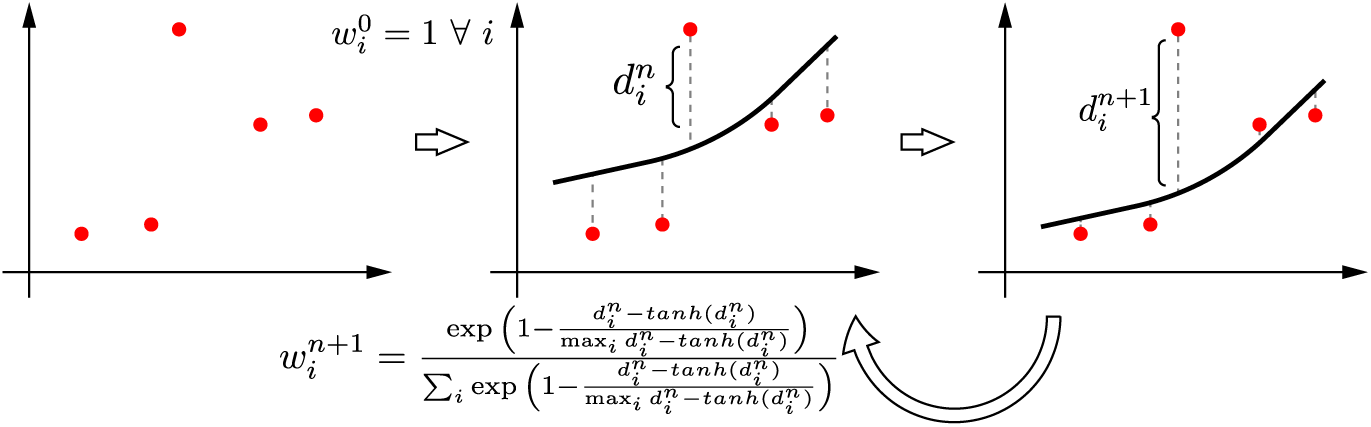
Iterative weighting technique for robust curve fitting.

The main idea is as follows. We maintain weights for all data points (*i*) in every iteration (*n*, starting from 0) as 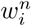. First, we fit a curve using the LM technique with weights of all data points as 1, thus 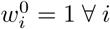. Second, we find the weight corresponding to every point for the next iteration 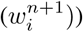 as:

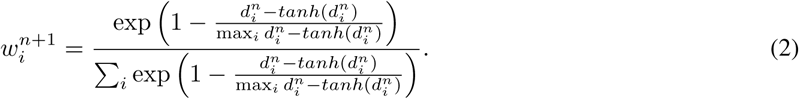

Simply, in the above equation, we first take *tanhshrink* function defined as *tanhshrink*(*x*) *= x − tanh*(*x*) for the distances of all points along *y* axis from the curve (*d_i_*). This is to have a higher value for points far from the curve and near 0 value for closer points. This, is then standardized by dividing with max value over all points and subtracted from 1 to get a weight corresponding to each point. This weight is then standardized using *softmax* function so that sum of all weights is 1. The curve is fit again using LM method, now with the new weights 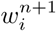. The algorithm converges when the sum total deviation of all weights becomes lower than a threshold value.

### Distribution Model Selection

To find the best fitting distribution model for the data corresponding to COVID-19, we studied the data on daily new confirmed COVID cases. Five sets of global data on daily new COVID-19 cases were used to fit parameters of different types of distributions. Finally, we identified the best performing 5 distributions. The results are shown in Table 1. We observe that using the iteratively weighted approach, the Inverse Weibull function fits the best to the COVID-19 dataset, as compared to the iterative versions of Gaussian, Beta (4-parameter), Fisher-Tippet (Extreme Value distribution), and Log Normal functions. When applied to the same dataset, Iterative Weibull showed an average MAPE of 12% lower than non-iteratively weighted Weibull. A step-by-step algorithm for iteratively weighted curve fitting using the GIW distribution (called “Robust Weibull”) is given in Algorithm 1.

**Table 1:**
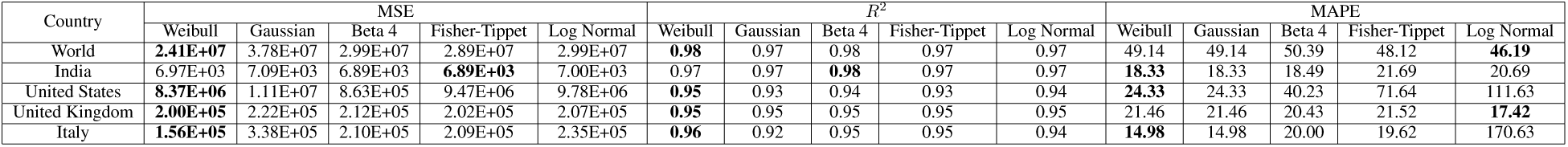
Preliminary Evaluation of different models. We observe that iterative fitting of Inverse Weibull performs significantly better than iterative fitting of other distributions like Gaussian, Beta (4-parameter), Fisher-Tippet (Extreme Value distribution), and Log Normal. The lowest value of MSE/MAPE and highest values of *R^2^* among all distributions are shown in bold.

### Analysis and Interpretation

To compare the proposed “Robust Weibull fitting” model, we use the baseline proposed by Jianxi Luo from SUTD ^3^. The comparison metrics include Mean Squared Error (MSE), Mean Absolute Percentage Error (MAPE) and Coefficient of determination (*R*^2^). Table 2 shows the model predictions of the spread of the COVID-19 for every major country for which sufficient data was available and model fits had *R*^2^ > 0.5 using the proposed model. As shown in the table, the proposed model performs significantly better than the baseline.

**Table 2:**
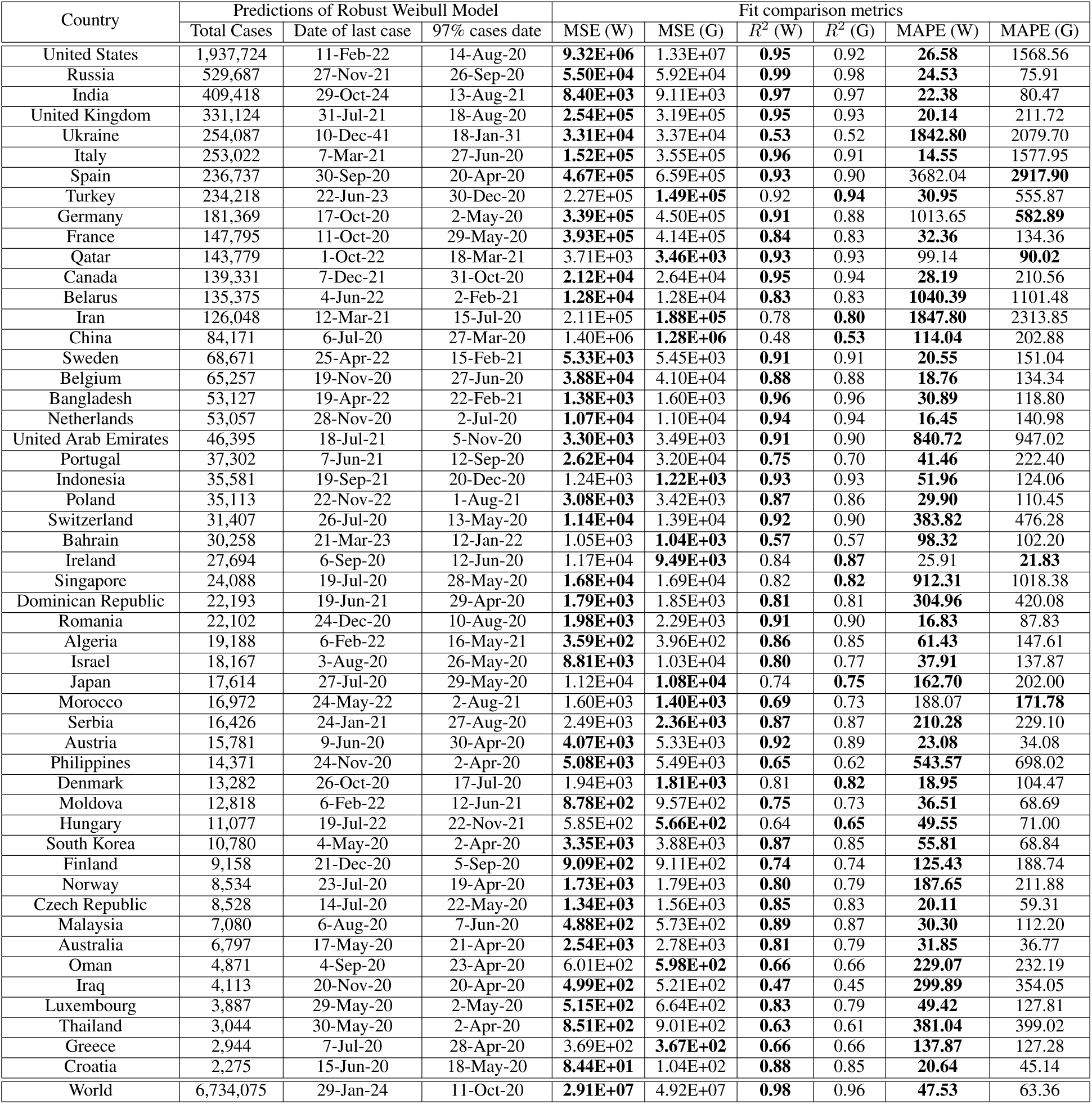
**Predictions and error comparisons**. Country wise predictions using Robust Weibull model and error comparison between Robust Weibull and baseline Gaussian Model. We predict the total number of cases that will be reached, and the last case date i.e. when the model predicts new cases < 1. We also predict the date when the total number will reach 97% of the total expected cases. Such data is critical to prepare the healthcare services in advance. The fit comparison metrics (with proposed model as *W* and baseline model as *G*) show that Mean Square Error (MSE) and the Mean Absolute Percentage Error (MAPE) of the proposed model are lower than baseline for most cases. The coefficient of determination (*R*^2^) is higher for the proposed model for most of the countries. The least MSE/MAPE and highest *R*^2^ values among the two models are shown in bold. Data upto 4 May, 2020 was used to create these results.

#### Algorithm 1 Robust Curve Fitting using Iterative weighting

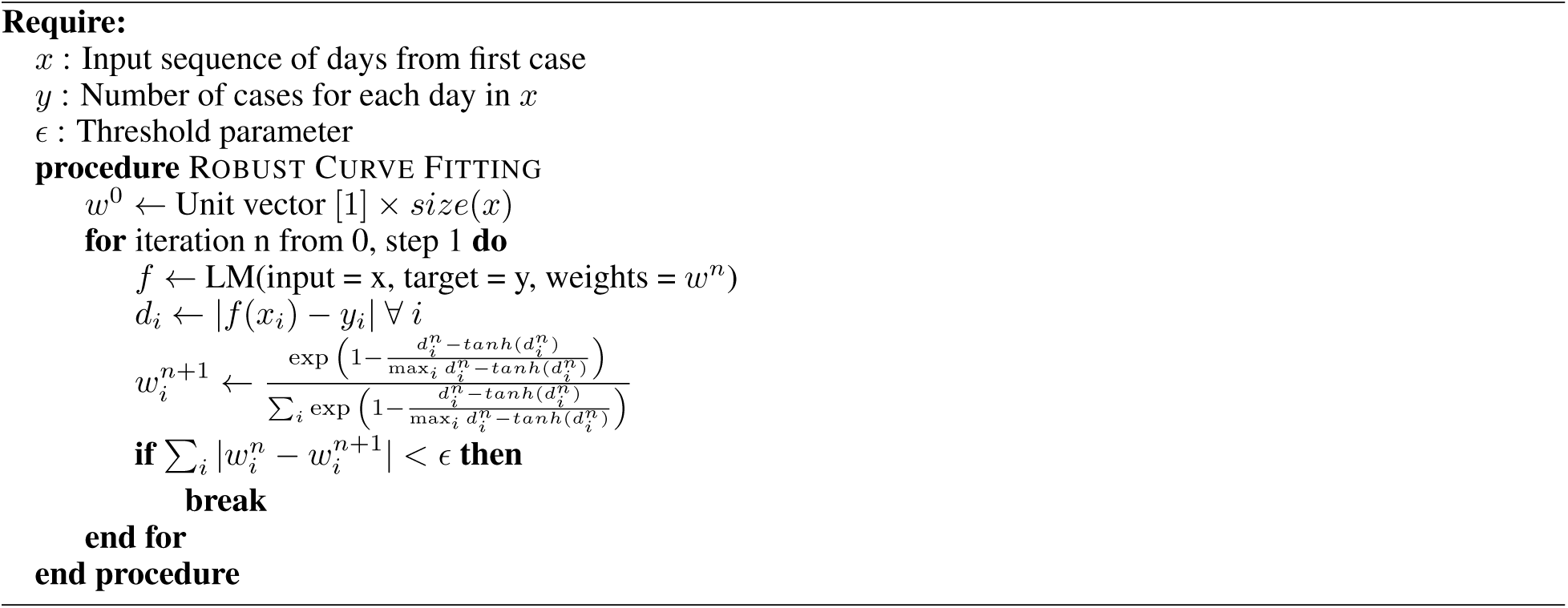

As shown in Figure 4^4^, the predictions of the baseline Gaussian model deployed by SuTD are overoptimistic. Following such models could lead to premature uplifting of the lockdown, causing adverse effect on management of the epidemic. Having better fit models, as proposed here, could help plan a better strategy, based on more accurate predictions and future scenarios.

**Figure 4:**
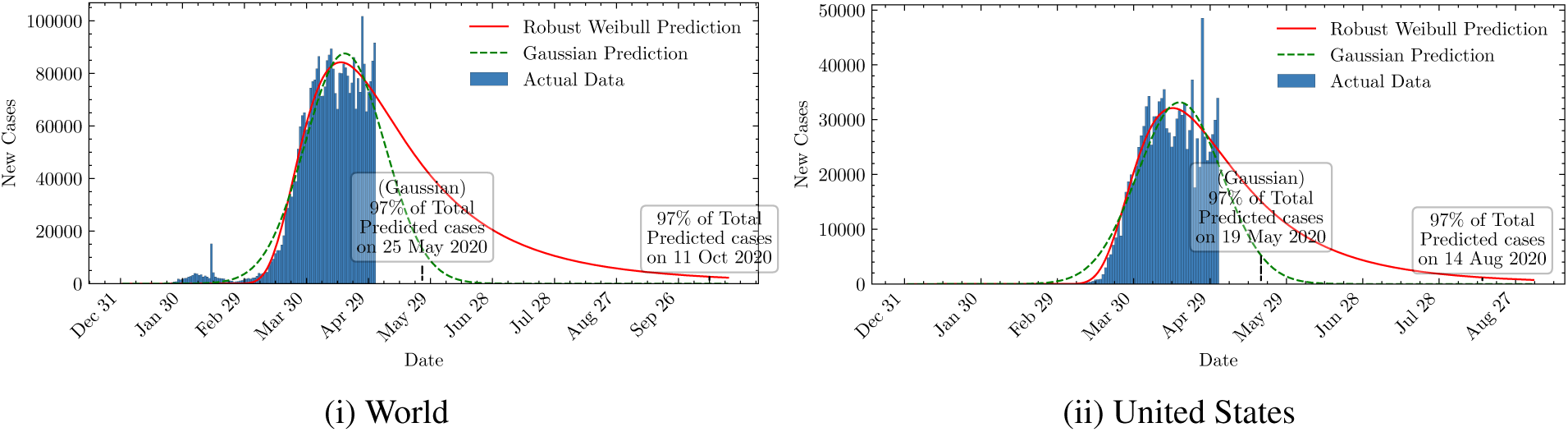
Comparison of predicted dates to reach 97% of the total expected cases by baseline Gaussian and proposed Robust Weibull models. The predicted end date of the pandemic in the baseline model are over-optimistic.

Figure 5 shows the total predicted number of cases for all countries across the globe. Here we have neglected those countries where the data is insufficient for making predictions, or the number of days for data is less than 30. As shown in Figure 4 explained in model section, the fit curve can be used to predict the number of cases that will have to be dealt by the country, assuming the same trend continues. The figure illustrates that the maximum number of total cases will be in the North America region. The number of cases will also be high in the European continent, Russia and eastern Asia, including china, the original epicenter of the disease.

**Figure 5:**
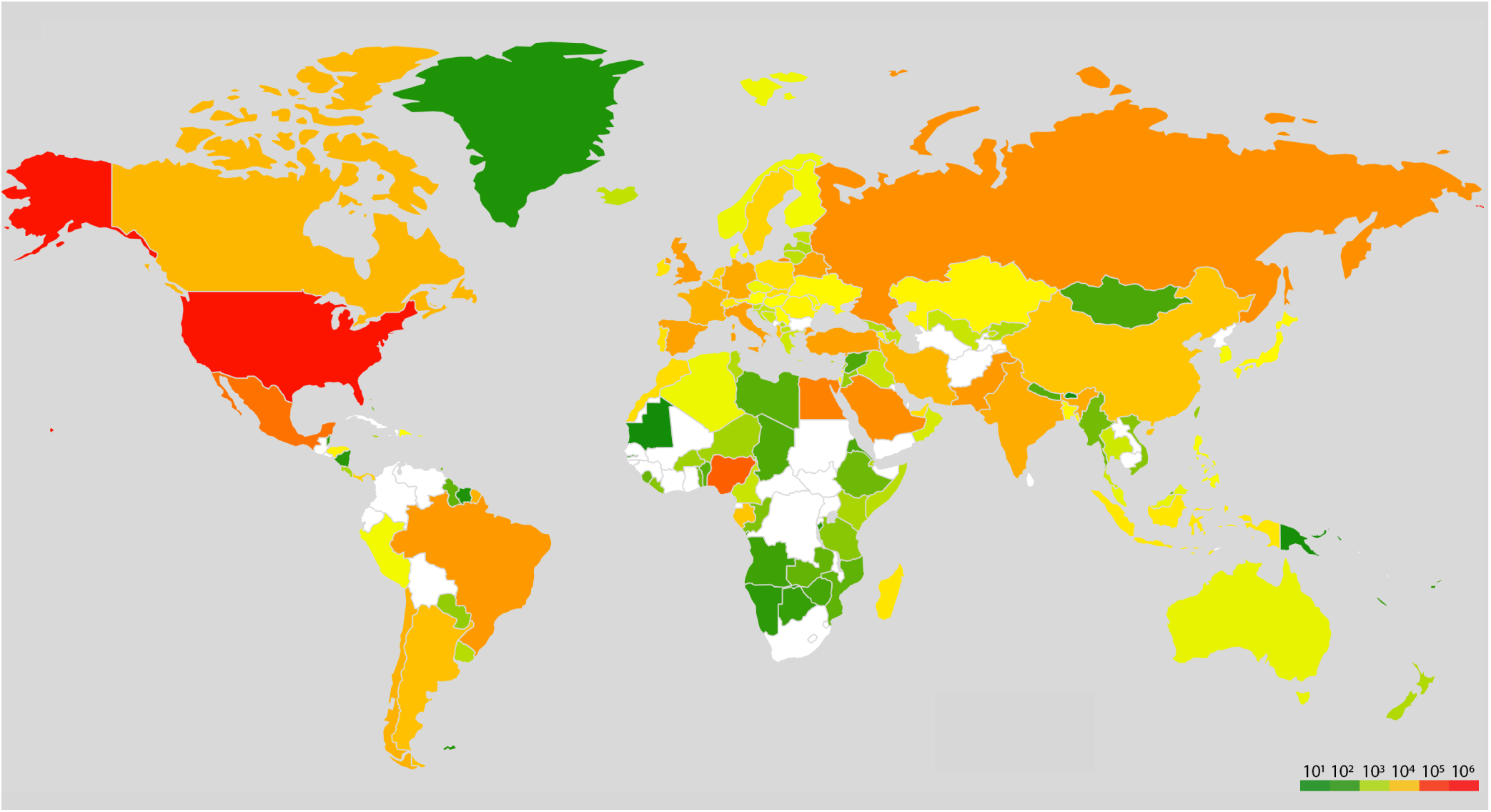
Global heat-map for total predicted cases for different countries as on May 4, 2020 (countries with insufficient data for prediction are shown in white)

## 3 Conclusions and Future Directions

In this study, we have discussed how improved mathematical modelling, Machine Learning and cloud computing can help to predict the growth of the epidemic proactively. Further, a case study has been presented which shows the severity of the spread of CoV-2 in countries worldwide. Using the proposed Robust Weibull model based on iterative weighting, we show that our model is able to make statistically better predictions than the baseline. The baseline Gaussian model shows an over-optimistic picture of the COVID-19 scenario. A poorly fitting model could lead to a non optimal decision making, leading to worsening of public health situation.

We propose the future directions as follows. Firstly, other important parameters like population density, distribution of age, individual and community movements, level of healthcare facilities available, strain type and virulence of the virus etc., need to be included in the regression model to further enhance the prediction accuracy. Secondly, models like ARIMA [27] can be integrated with Weibull function for further time series analysis and predictions. Thirdly, ML can be utilized to predict the structure and function of various proteins associated with CoV-2 and their interaction with the host human proteins and cellular environment. The contribution of various socio-economic variables that determine the vulnerability, spread and progression of the epidemic can be predicted by developing suitable algorithms. AI based proactive measures can be taken to prevent the spread of the virus to sensitive groups in the society. Real time sensors can be used, for example in traffic camera or surveillance, which track COVID-19 symptoms based on visual imaging and tracking Apps, and inform respective hospitals and administrative authorities for punitive action [28]. Tracking needs to cover all stages from ports of entries to public places and hospitals [29]. The research directions and challenges are summarized in Figure 6.

**Figure 6:**
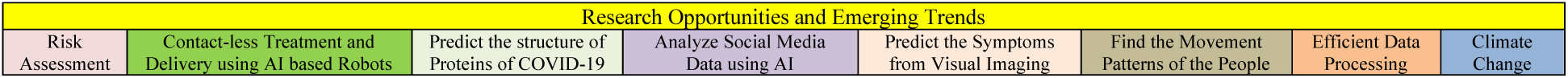
Future Research Directions and Open Challenges

## 4 Software Availability

Our prediction model is available online at https://github.com/shreshthtuli/covid-19-prediction. The dataset used for this work is the *Our World Dataset*, available at https://github.com/owid/covid-19-data/tree/master/public/data/. Few interactive graphs can be seen at https://collaboration.coraltele.com/covid/.

## Data Availability

Our prediction model is available online at https://github.com/shreshthtuli/covid-19-prediction.
The dataset used for this work is the Our World Dataset, available at https://github.com/owid/covid-19-data/tree/master/public/data/. Few interactive graphs can be seen at https://collaboration.coraltele.com/covid/.

## Appendix: Real data from WHO with predicted curves of proposed and baseline models

All data uptil 4 May 2020 has been used to generate results.

**Figure 7:**
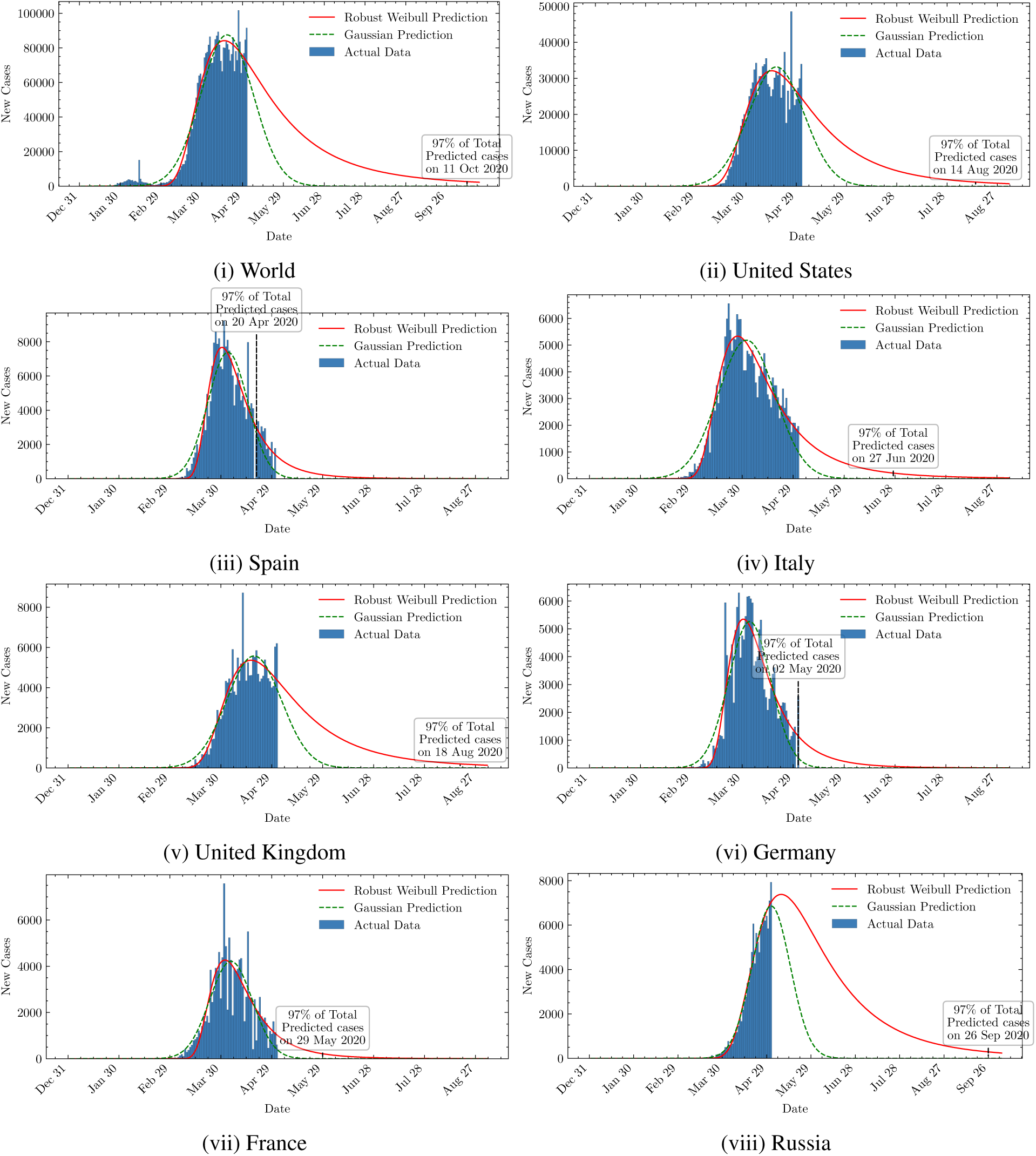
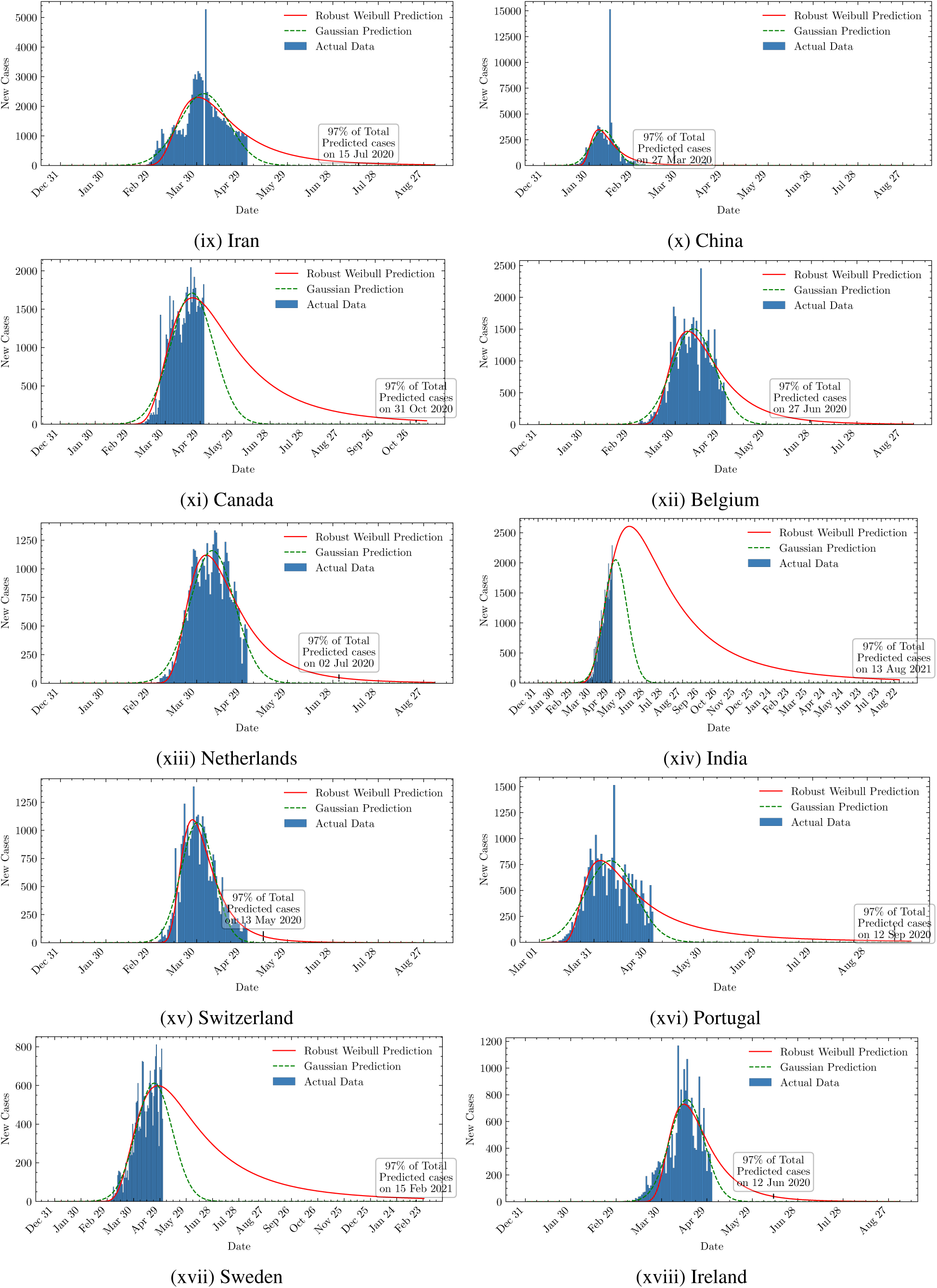
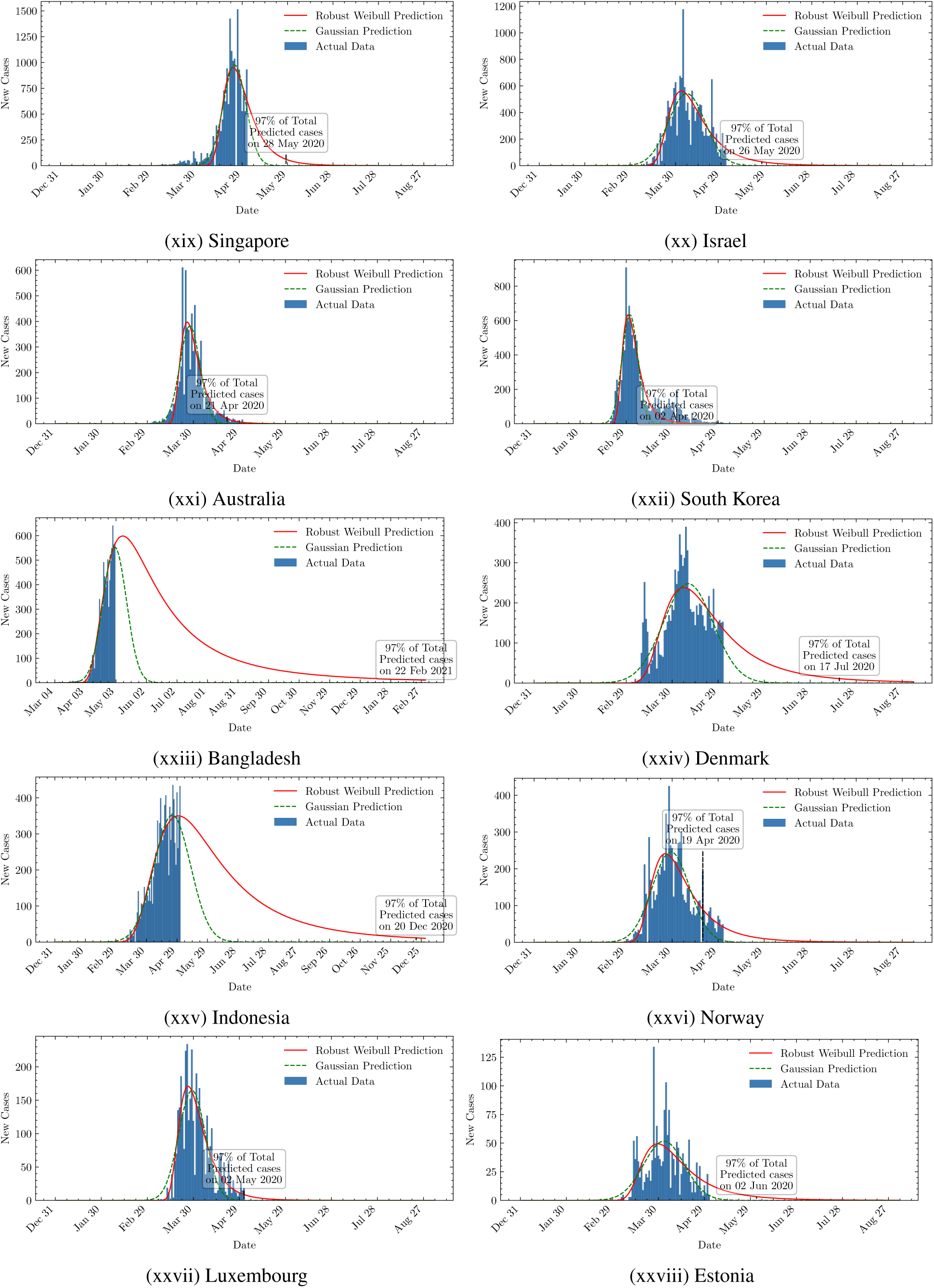
New cases for different countries

### Author’s Biography

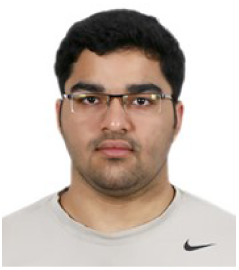

**Shreshth Tuli** is an undergraduate student at the Department of Computer Science and Engineering at Indian Institute of Technology - Delhi, India. He is also a co-founder of Qubit Inc. company which works on providing next generation solutions for industrial problems. He is a national level Kishore Vaigyanic Protsahan Yojana (KVPY) scholarship holder for excellence in science and innovation. He has worked as a visiting researcher at the Cloud Computing and Distributed Systems (CLOUDS) Laboratory, Department of Computing and Information Systems, the University of Melbourne, Australia. His research interests include Internet of Things (IoT), Fog Computing, Network Design, and Artificial Intelligence.

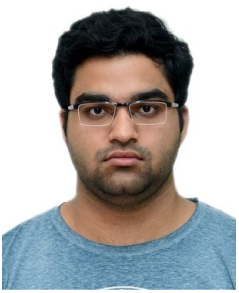

**Shikhar Tuli** is an undergraduate student at the Department of Electrical Engineering at Indian Institute of Technology - Delhi, India. He is the founder and CEO of Qubit Inc. He has worked remotely with the Cloud Computing and Distributed Systems (CLOUDS) Laboratory, Department of Computing and Information Systems, the University of Melbourne, Australia in the realization of the FogBus framework. He has also worked at the Embedded Systems Laboratory, EPFL, Switzerland in the design of low-power and physics-optimized Edge devices made from emerging Non-Volatile Memories. His research interests include Internet of Things (IoT), In-memory and Neuromorphic computing architectures and Nanoelectronics. He specializes in designing novel hardware technologies that are valuable to both industry and academia.

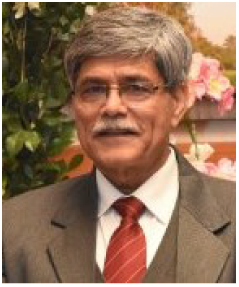

**Rakesh Tuli** is Senior Research Advisor; J C Bose National Fellow, UIET, Panjab University, Chandigarh. Before this, he was executive Director, National Agri-Food Biotech Institute, Mohali; and Director, National Botanical Research Institute, Lucknow. He has many publications in reputed journals and conferences including Nature Biotechnology. His research includes Genomics and Transgenic Approaches to Improving Plants for Agricultural and Health/Medicinal Applications.

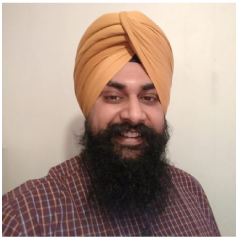

**Sukhpal Singh Gill** is a Lecturer (Assistant Professor) in Cloud Computing at School of Electronic Engineering and Computer Science, Queen Mary University of London, UK. Prior to this, Dr. Gill has held positions as a Research Associate at the School of Computing and Communications, Lancaster University, UK and also as a Postdoctoral Research Fellow at CLOUDS Laboratory, The University of Melbourne, Australia. His research interests include Cloud Computing, Fog Computing, Software Engineering, Internet of Things and Big Data.

## Footnotes

0 Abbreviations: ML, Machine Learning; SARS-CoV-2; Severe Acute Respiratory Syndrome Coronavirus 2, COVID-19, Coronavirus disease

1 Our World In Data: COVID-19 Dataset; source: https://github.com/owid/covid-19-data/tree/master/public/data/

2 Situation Reports: WHO; source: https://www.who.int/emergencies/diseases/novel-coronavirus-2019/situation-reports

1 Azure Cloud VMs: https://azure.microsoft.com/en-au/pricing/calculator/

3 When Will COVID-19 End, DDI Lab, SUTD: https://ddi.sutd.edu.sg/when-will-covid-19-end

4 Curves and predictions of all countries have been given in Appendix

